# Is energy density an implicit part of the purpose behind ultra-processed food? A comparison of relative energy density across ultra-processed and minimally processed food

**DOI:** 10.1101/2025.10.21.25338444

**Authors:** Samuel J. Dicken, Adrian Brown

## Abstract

**Introduction:** Diets high in ultra-processed food (UPF) are linked with greater energy intake and less favourable weight change than diets high in minimally processed food (MPF), as defined by the Nova classification. Energy density is an important factor influencing palatability and energy intake and suggested as a key mechanism of UPF. Whether there are consistent differences in energy density between MPF and UPF across different types of food and drink is unknown.

**Methods:** This study compared the energy density of like-for-like MPF and UPF within food and drink groups from the nationally representative UK National Diet and Nutrition Survey (NDNS) Intake24 food and drink database, as well as within ‘healthy’ items only (containing no red front-of-package label traffic lights).

**Results:** Overall, UPF food had a significantly higher energy density than MPF food (1.03 kcal/g vs. 2.07 kcal/g; p < 0.001). Most UPF (breakfast cereals, puddings, yogurt, fromage frais and other dairy desserts, egg end egg dishes, meat, fish and meat/fish dishes, fruit and vegetables, and potato and potato products) had a significantly higher energy density than MPF from the same food group. Overall, UPF drinks had a significantly lower energy density than MPF drinks (0.20 kcal/g vs. 0.39 kcal/g; p = 0.003). Findings were similar when assessing healthy food and drink only.

**Conclusion:** These results suggest that energy density may be an implicit part of UPF and have implications for understanding the role of energy density in the context of the purpose of food processing. These findings call for caution in over adjusting for energy density in studies of UPF.

## Introduction

Mounting evidence links high intakes of ultra-processed food (UPF) (multi-ingredient formulations designed with the primary purpose of maximising profit)^1^ with poor health outcomes, including obesity, obesity-related disease and premature mortality^1,2^. In particular, UPF may promote greater energy consumption and unfavourable weight change compared with minimally processed food (MPF)^1^, as evidenced across several clinical trials^3–5^. Several mechanisms are proposed to explain this difference, including energy density, texture and hyperpalatability^4,6^.

In particular, energy density has a strong influence on total energy intake^7^. Covert manipulation to increase or decrease energy density in single meals or whole diets has been shown to increase or decrease total energy intake, respectively^8,9^. And across UPF and MPF, the energy density of food and drink can vary greatly^10^. As a result, there have been arguments that analyses comparing UPF versus non-UPF on health outcomes should control for energy density^11^. However, given its role in palatability and food choice, energy density is likely to be implicated in the purpose of ultra-processing in creating ultra-profitable foods^12,13^. Indeed, evidence to date suggests UPF are more energy dense than MPF in the US^14^ and UK^10^. Furthermore, UPF without high levels of fat, saturated fat, salt and/or sugar (and considered ‘healthy’) are still more energy dense than MPF^10^. This points towards a potential innate role of energy density in UPF.

Importantly, small differences in energy density can significantly influence energy intake^8^. With wide variations in energy density observed across different types of food and drink, even in those considered healthy^10^, this has the potential to impact on body weight and health-related outcomes. But no study to date has examined whether there are consistent differences in energy density across similar types of food and drink, that differ in purpose of processing (i.e. MPF vs UPF). The one previous UK study assessing differences in energy density between MPF and UPF compared across all items, or ‘healthy’ items only^10^. Therefore, it is important to determine whether differences in energy density exist between MPF and UPF after accounting for type of food or drink and nutritional quality, to better define the role of energy density as a confounder or mediator in studies of UPF.

The aim of this study was to assess differences in energy density between MPF and UPF, first across all items within food and drink groups, and second, across only ‘healthy’ items within food and drink groups.

## Methods

### Data sources

Details on the methods used have been reported elsewhere^10^. Briefly, the nutrient composition data of food and drink items from Year 12 of the UK National Diet and Nutrition Survey (NDNS) was used^15^. The nutritional values in the nutrient databank are determined from multiple sources, mainly from the UK Composition of Foods Integrated Dataset^16^, which is complemented by manufacturer data from food labels and the web, and the Food Standards Agency (FSA) Food Recipes Database^17^.

### Nova classification

The process of coding NDNS food and drink items into the Nova classification is described previously^10^. Of the 3105 items in the database, 55.4% were UPF (n = 1650) and 33.1% were MPF (n *=* 986). This analysis was restricted to MPF and UPF, as processed culinary ingredients (PCI) are not consumed in isolation, and due to limited numbers of processed food (PF) within food and drink groups for comparison.

### Front-of-package label traffic light classification

The process of coding NDNS food and drink items into front-of-package label multiple traffic lights (FOPL MTL) according to the Department of Health and Food Standards Agency guidance for fat, saturated fat, total sugar and salt content^18^, is described previously^10,19^. Items were classified as ‘healthy’ or ‘unhealthy’, based on the presence or absence of a red FOPL traffic light for fat, saturated fat, total sugar or salt, as conducted previously^10,19^.

### Food and drink groups

Food and drink groups were defined by NDNS in the nutrient databank^15^. Food and drink groups spanning both MPF and UPF were included (Supplementary Table 1). Similar food and drink groups were also combined into broader groups to allow comparison between MPF and UPF (Supplementary Table 1). Food and drink groups not spanning MPF and UPF (e.g. containing 2 or fewer MPF) were excluded from the analysis due to limited comparability (Supplementary Table 1). No items were excluded from food and drink groups (except infant formula and creams from milk, to ensure comparability).

### Statistical analysis

Non-parametrically distributed variables were described using medians and interquartile ranges (IQR), and categorical variables using counts and percentages. Comparisons of energy density (kcal/g) between MPF and UPF across food and drink groups were analysed using Mann-Whitney U tests. Statistical significance was set at p < 0.05 and not adjusted for multiple comparisons. Analyses were conducted in R (V.2024.12.1+563).

## Results

### Overall

A total of 934 (851 food items) MPF and 846 (694 food items) UPF were included in the analysis. The energy density of MPF and UPF food and drink overall are reported in Table 1 and Figure 1. The energy density of MPF and UPF food and drink excluded from the analysis are reported in Supplementary Table 2.

**Table 1.**
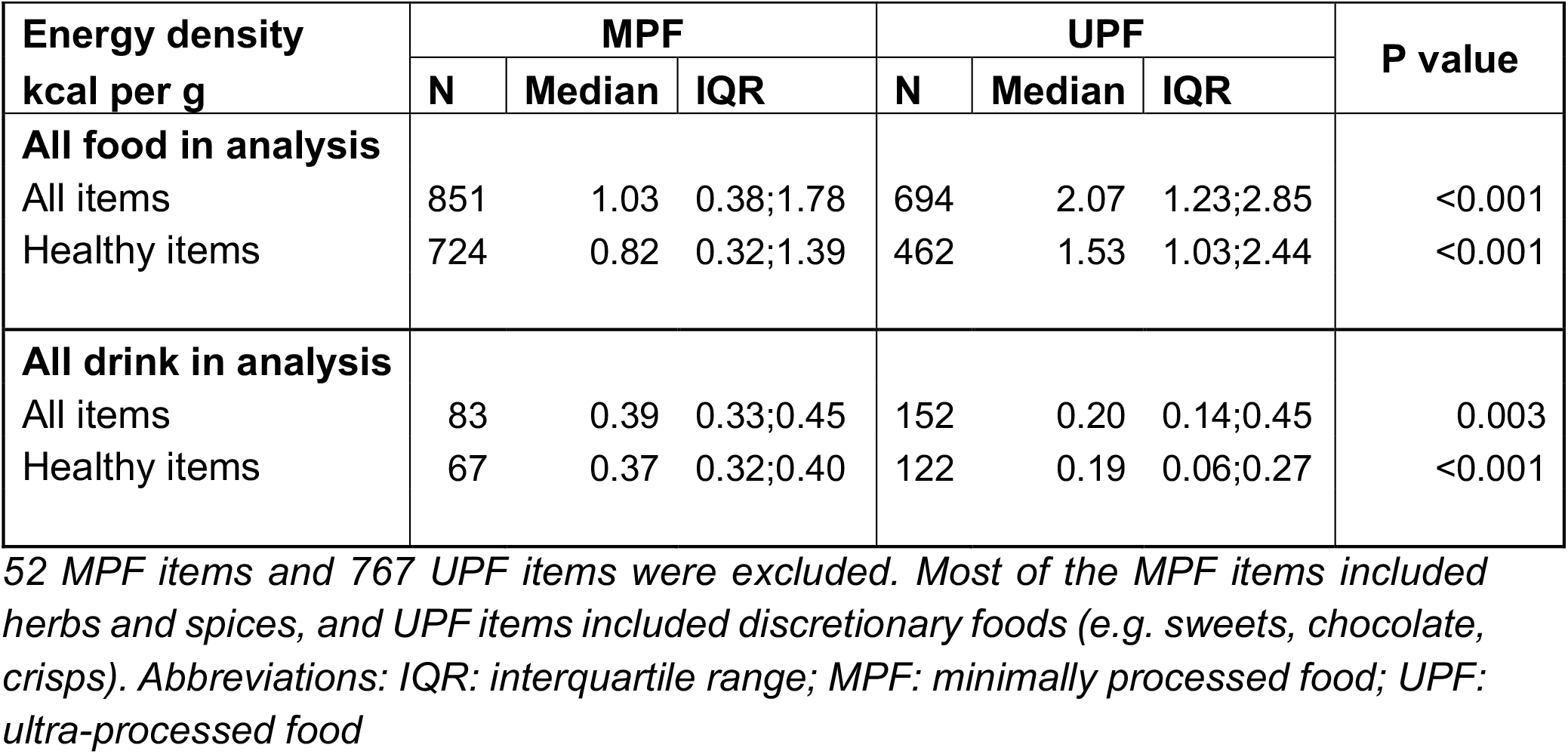
Energy density of minimally processed and ultra-processed food and drink, overall, and within healthy items only.

| Energy density<br>kcal per g | MPF |  |  | UPF |  |  | P value |
| --- | --- | --- | --- | --- | --- | --- | --- |
|  | N | Median | IQR | N | Median | IQR |  |
| **All food in analysis** |  |  |  |  |  |  |  |
| All items | 851 | 1.03 | 0.38;1.78 | 694 | 2.07 | 1.23;2.85 | <0.001 |
| Healthy items | 724 | 0.82 | 0.32;1.39 | 462 | 1.53 | 1.03;2.44 | <0.001 |
| **All drink in analysis** |  |  |  |  |  |  |  |
| All items | 83 | 0.39 | 0.33;0.45 | 152 | 0.20 | 0.14;0.45 | 0.003 |
| Healthy items | 67 | 0.37 | 0.32;0.40 | 122 | 0.19 | 0.06;0.27 | <0.001 |

52 MPF items and 767 UPF items were excluded. Most of the MPF items included herbs and spices, and UPF items included discretionary foods (e.g. sweets, chocolate, crisps). Abbreviations: IQR: interquartile range; MPF: minimally processed food; UPF: ultra-processed food

**Figure 1:**
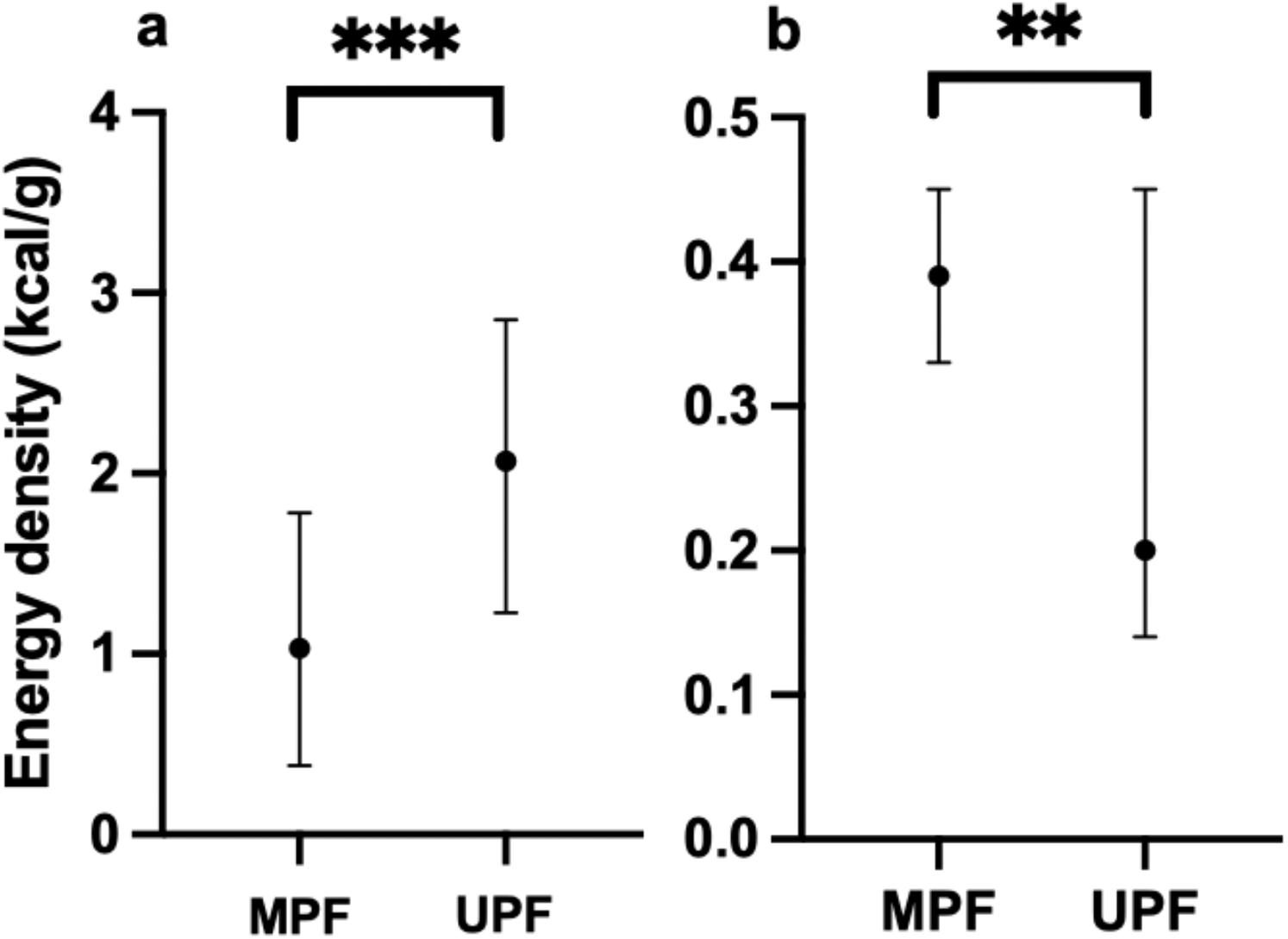
Energy density of minimally processed and ultra-processed food and drink overall. *a: food; b: drink. Abbreviations: MPF: minimally processed food; UPF: ultra-processed food.*

Overall, MPF food had a significantly lower energy density than UPF food (MPF: 1.03 kcal/g (IQR: 0.38, 1.78); UPF: 2.07 kcal/g (IQR: 1.23, 2.85), p < 0.001). Healthy MPF food also had a significantly lower energy density than healthy UPF food (MPF: 0.82 kcal/g (IQR: 0.32, 1.39); UPF: 1.53 kcal/g (IQR: 1.03, 2.44), p < 0.001).

Overall, MPF drinks had a significantly higher energy density than UPF drinks (MPF: 0.39 kcal/g (IQR: 0.33, 0.45); UPF: 0.20 kcal/g (IQR: 0.14, 0.45), p = 0.003). Healthy MPF drinks also had a significantly higher energy density than healthy UPF drinks (MPF: 0.37 kcal/g (IQR: 0.32, 0.40); UPF: 0.19 kcal/g (IQR: 0.06, 0.27), p < 0.001).

### Food

The energy density of MPF and UPF food groups are reported in Table 2.

**Table 2.**
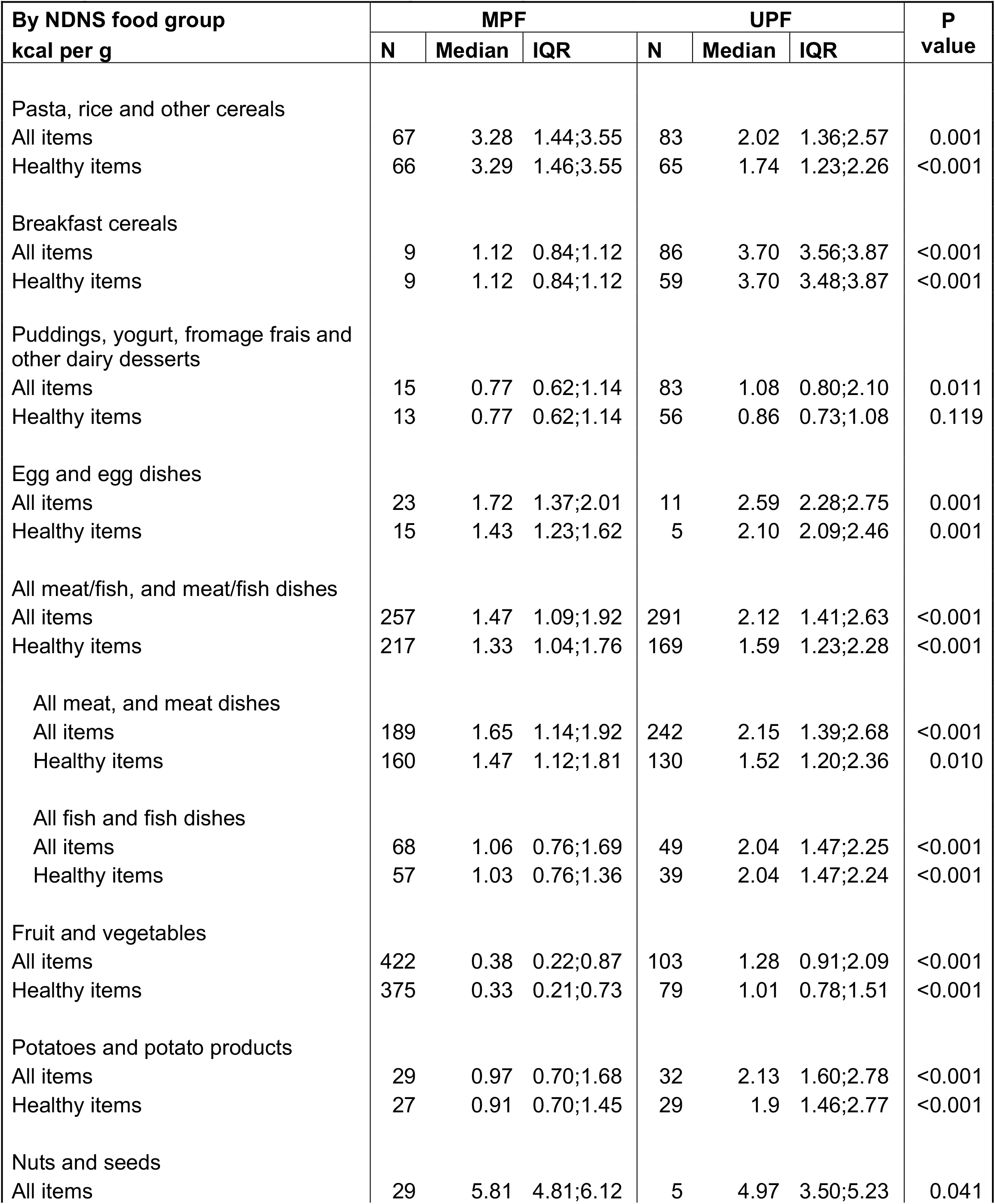
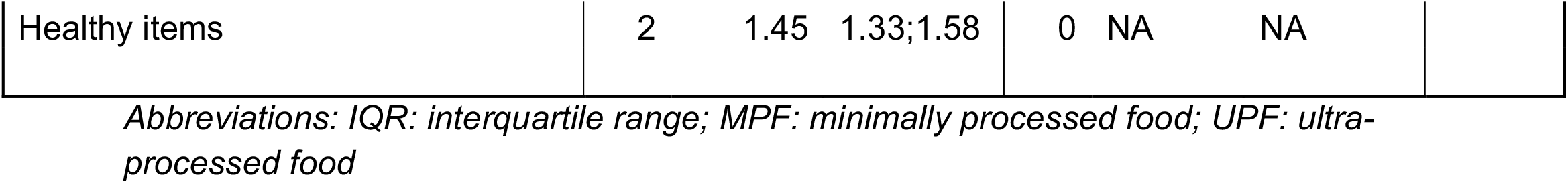
Energy density of minimally processed and ultra-processed food by group, overall, and within healthy items only.

Overall, MPF breakfast cereals (MPF: 1.12 kcal/g (IQR: 0.84, 1.12); UPF: 3.70 kcal/g (IQR: 3.56, 3.87), p < 0.001), puddings, yogurt, fromage frais and other dairy desserts (MPF: 0.77 kcal/g (IQR: 0.62, 1.14); UPF: 1.08 kcal/g (IQR: 0.80, 2.10), p = 0.011), egg end egg dishes (MPF: 1.72 kcal/g (IQR: 1.37, 2.01); UPF: 2.59 kcal/g (IQR: 2.28, 2.75 ), p = 0.001), meat, fish and meat/fish dishes (MPF: 1.47 kcal/g (IQR: 1.09, 1.92); UPF: 2.12 kcal/g (IQR: 1.41, 2.63), p < 0.001), fruit and vegetables (MPF: 0.38 kcal/g (IQR: 0.22, 0.87); UPF: 1.28 kcal/g (IQR: 0.91, 2.09), p < 0.001), and potato and potato products (MPF: 0.97 kcal/g (IQR: 0.70, 1.68); UPF: 2.13 kcal/g (IQR: 1.60, 2.78), p < 0.001) had a significantly lower energy density than UPF from the same food group. Overall, MPF nuts and seeds (MPF: 5.81 kcal/g (IQR: 4.81, 6.12); UPF: 4.97 kcal/g (IQR: 3.50, 5.23), p = 0.041), and pasta, rice and other cereals (MPF: 3.28 kcal/g (IQR: 1.44, 3.55); UPF: 2.02 kcal/g (IQR: 1.36, 2.57), p = 0.001) had a significantly higher energy density than UPF from the same food group.

Results were similar when comparing healthy food items only, except the energy density of healthy puddings, yogurt, fromage frais and other dairy desserts did not significantly differ between MPF and UPF.

The energy density of MPF and UPF groups combined within the broader groups are reported in Supplementary Table 3.

Across excluded food, MPF were significantly less energy dense than UPF, both overall (MPF: 2.58 kcal/g (IQR: 0.72, 3.23); UPF: 3.40 kcal/g (IQR: 2.31, 4.36), p <0.001), and within healthy items only (MPF: 0.74 kcal/g (IQR: 0.26, 2.52); UPF: 2.30 kcal/g (IQR: 1.46, 2.74), p < 0.001) (Supplementary Table 2).

### Drink

The energy density of MPF and UPF drink groups are reported in Table 3.

**Table 3.**
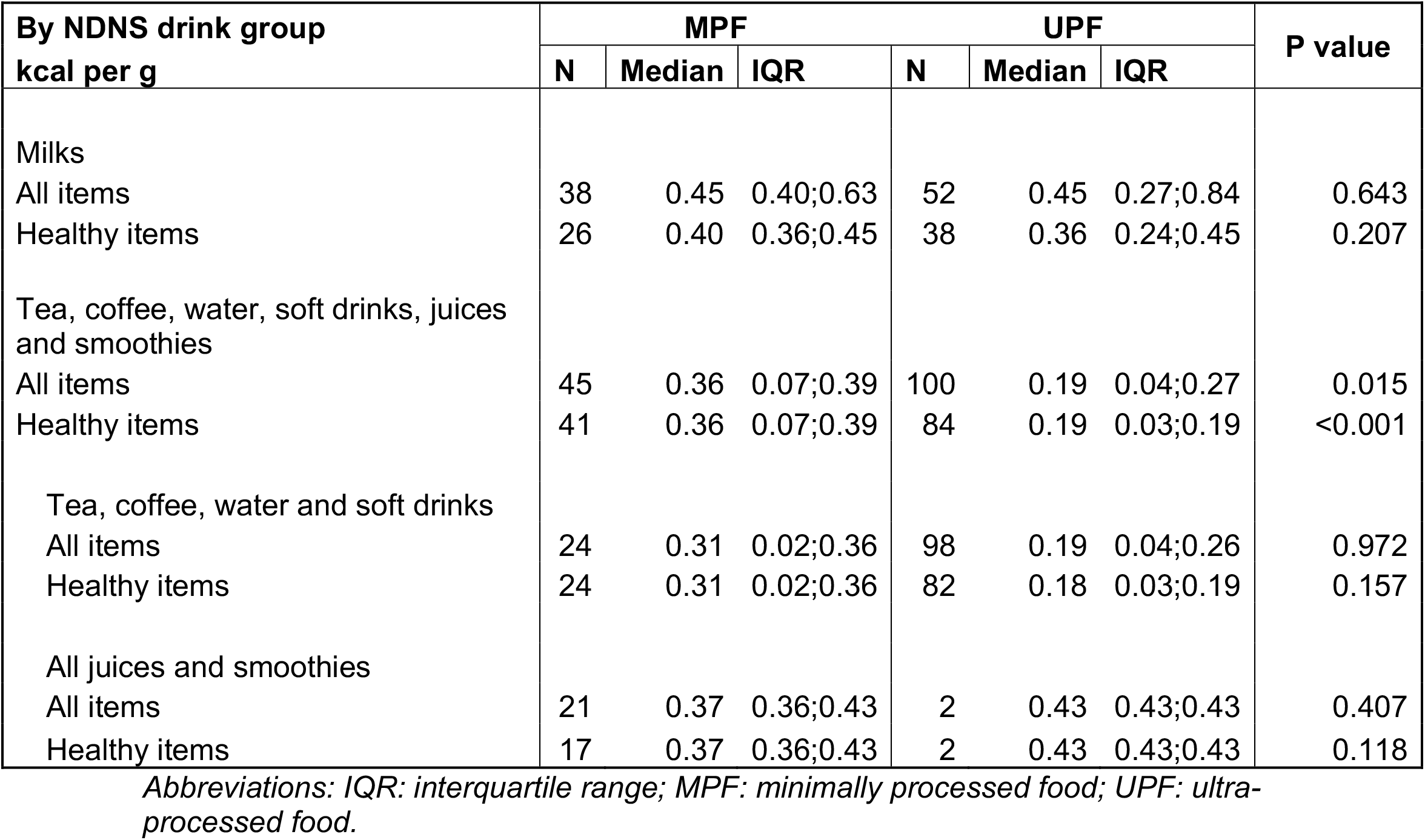
Energy density of minimally processed and ultra-processed drink by group, overall, and within healthy items only.

| By NDNS drink group<br>kcal per g | MPF |  |  | UPF |  |  | P value |
| --- | --- | --- | --- | --- | --- | --- | --- |
|  | N | Median | IQR | N | Median | IQR |  |
| Milks |  |  |  |  |  |  |  |
| All items | 38 | 0.45 | 0.40;0.63 | 52 | 0.45 | 0.27;0.84 | 0.643 |
| Healthy items | 26 | 0.40 | 0.36;0.45 | 38 | 0.36 | 0.24;0.45 | 0.207 |
| Tea, coffee, water, soft drinks, juices<br>and smoothies |  |  |  |  |  |  |  |
| All items | 45 | 0.36 | 0.07;0.39 | 100 | 0.19 | 0.04;0.27 | 0.015 |
| Healthy items | 41 | 0.36 | 0.07;0.39 | 84 | 0.19 | 0.03;0.19 | <0.001 |
| Tea, coffee, water and soft drinks |  |  |  |  |  |  |  |
| All items | 24 | 0.31 | 0.02;0.36 | 98 | 0.19 | 0.04;0.26 | 0.972 |
| Healthy items | 24 | 0.31 | 0.02;0.36 | 82 | 0.18 | 0.03;0.19 | 0.157 |
| All juices and smoothies |  |  |  |  |  |  |  |
| All items | 21 | 0.37 | 0.36;0.43 | 2 | 0.43 | 0.43;0.43 | 0.407 |
| Healthy items | 17 | 0.37 | 0.36;0.43 | 2 | 0.43 | 0.43;0.43 | 0.118 |

*Abbreviations: IQR: interquartile range; MPF: minimally processed food; UPF: ultra-* *processed food.*

The energy density of MPF and UPF milk did not significantly differ overall, or within healthy drinks only. UPF juices and smoothies, and tea, coffee, water and soft drinks were significantly less energy dense than comparable MPF, both overall (MPF: 0.36 kcal/g (IQR: 0.07, 0.39); UPF: 0.19 kcal/g (IQR: 0.04, 0.27), p = 0.015), and within healthy items only (MPF: 0.36 kcal/g (IQR: 0.07, 0.39); UPF: 0.19 kcal/g (IQR: 0.03, 0.19), p < 0.001). There were no significant differences between MPF and UPF for tea, coffee, water, or soft drinks, either overall or within the healthy drinks category.

## Discussion

This is the first study to compare differences in energy density between MPF and UPF across different types of food and drink in a nationally representative UK food and drink database, and by nutritional quality. MPF food had a significantly lower energy density than UPF food, both overall and for healthy items only. For drinks, the opposite was observed. MPF drinks had a significantly higher energy density than UPF drinks,both overall and for healthy items only. However, energy density within each specific drink group did not significantly differ between MPF and UPF.

Calls to control for energy density^11^ have typically relied on arguments from analogy, that some UPF are lower in energy density than some MPF (e.g. artificially-sweetened beverages versus whole milk). However, UPF are generally designed to be highly attractive and palatable in order to displace MPF and outcompete rival UPF products^1^. A key part of this is acting on the innate human preference for energy dense foods^13^. Arguments to control for energy density have not considered the necessary comparison of energy density between like-for-like MPF/UPF food and drink. The previous UK analysis found that healthy MPF (food and drink) had an average energy density of 0.75 kcal/g (IQR: 0.32, 1.34), compared with 1.08 kcal/g (IQR: 0.59, 1.75) for PF, and 1.52 kcal/g (IQR: 0.77, 2.43) for UPF. This analysis moves away from binary views of UPF as a uniform group, and towards nuanced perspectives on how the purpose of ultra-processing influences sensory properties and food-level characteristics compared with like-for-like MPF. In this regard, after accounting for type of food and nutritional quality, this study found significant differences in energy density between MPF and UPF, consistent across several food groups. The results here support the role of energy density being implicated as a core aspect of UPF.

Small differences in energy density, as observed here across a number of key food groups between MPF and UPF, could result in significant changes in energy intake^8^. This is particularly important for non-beverage energy density^6^. These small average increases in energy density of UPF compared with MPF within the same types of food, when scaled up across the whole diet, may have meaningful impacts on energy intake and body weight. These results have implications for understanding the role of energy density in the context of UPF and potentially calls for caution in over adjusting for energy density in studies of UPF to determine the real-world influence on health (i.e. as a mediator versus confounder).

Further research needs to explore possible differences in wider food-level characteristics and sensory properties linked with excess intake between UPF and comparable non-UPF, such as textural changes or portion sizes, alongside energy density. Importantly, in the UK, there is currently no dietary guidance on energy density or sensory properties, or public health policy explicitly focussed on energy density or sensory properties. Therefore, understanding which food-level characteristics are shaped by this purpose of profit maximisation can help to better identify key mechanisms and identify relevant upstream policy actions on UPF.

### Strengths

There are several strengths of this current analysis, which include the use of a nationally representative UK food and drink database, comparing MPF and UPF using UK government defined groupings of food and drink, and comparing MPF and UPF based on their nutritional quality according to UK government MTL FOPL.

### Limitations

Limitations include that the NDNS database contains averages of items based on several example items of that food or drink. Therefore, not every MPF or UPF item available in the UK was compared. Processed food was not included in the analysis due to low numbers across food and drink groups. The energy density of all items within each included food/drink group was compared as reported in the database, and no items were excluded from food/drink groups (except excluding formula and creams in milk). However, the energy density of consumed foods following preparation may differ (e.g. breakfast cereals with added milk, uncooked minimally processed flours).

## Conclusions

In a nationally representative database of food and drink in the UK, different types of UPF food have a significantly higher energy density than comparable MPF food, and different types of UPF drinks have a significantly lower energy density than comparable MPF drinks. These significant differences remain when comparing healthy food and drink items only. These results have implications for understanding the role of energy density in the context of the purpose of food processing and calls for caution in over adjusting for energy density in studies of UPF.

## Acknowledgements

The authors would like to thank the Intake24 team for their administrative support, Fernanda Rauber and her team for sharing their full NOVA classification of food and drink items from NDNS rolling programme Years 1–11.

## Ethics

No humans or animals were involved in this study. This study did not require ethical approval as it was an analysis of a food and drink database.

## Author contributions

SD: conceptualisation, data analysis, first manuscript draft. AB: data interpretation, review of manuscript.

## Data Availability Statement

Data can be made available upon request. For access to Intake24 datasets, please contact the Intake24 team; https://intake24.co.uk.

