## Supplementary Materials for "Is energy density an implicit part of the purpose behind ultra-processed food? A comparison of relative energy density across ultra-processed and minimally processed food"

**Supplementary Table 1. Included and excluded food and drink groups from the analysis, and grouping of included groups.**

**Supplementary Table 2. Energy density of excluded minimally processed and ultra-processed food and drink, overall, and within healthy items only.**

**Supplementary Table 3. Energy density of minimally processed and ultra-processed food by group, overall, and within healthy items only.**

**Supplementary Table 1. Included and excluded food and drink groups from the analysis, and grouping of included groups*.**

| **Included Food and Drink Groups** | | **Excluded food and drink groups** |
| --- | --- | --- |
| **Food** | **Drink** |  |
| **Pasta, rice and other cereals** | **All milks** | Breads (2,3,4, 59) |
| #1 Pasta, rice and other cereals (including pizza) | #10 Whole milk | Biscuits (7) |
|  | #11 Semi-skimmed milk | Buns, cakes, pastries and fruit pies (8) |
| **All breakfast cereals** | #12 Skimmed milk | Other milk (infant formula and creams) (13A & 13B) |
| #5 High fibre breakfast cereals | #13R Other milk (excluding formula and creams) | Cheese (14) |
| #6 Other breakfast cereals | #60 1% fat milk | Butter, margarine, spreads (17-21) |
|  |  | Sugars, preserves and sweet spreads (41) |
| **Yogurt and fromage frais** | **Tea, coffee, water, soft drinks, juices and smoothies** | Crisps and savoury snacks (42) |
| #15 Yogurt, fromage frais and other dairy desserts | #45 Fruit juice | Sugar confectionery (43) |
|  | #61 Smoothies 100% fruit and/or juice | Chocolate confectionery (44) |
| **Puddings, Yogurt, fromage frais and other dairy desserts** | #51 Tea, coffee and water | Miscellaneous (50) |
| #9 Puddings | #58 Soft drinks, not low calorie | Commercial toddlers’ foods and drinks (52) |
| #15 Yogurt, fromage frais and other dairy desserts | #59 Soft drinks, low calorie | Ice cream (53) |
|  |  | Dietary supplements (54) |
| **Egg and egg dishes** | **All juices and smoothies** | Artificial sweeteners (55) |
| #16 Egg and egg dishes | #45 Fruit juice | Sandwiches (62) |
|  | #61 Smoothies 100% fruit and/or juice | Alcohol (47,48,49) |
| **All meat/fish, and meat/fish dishes (22-35)** |  |  |
| #22 Bacon and ham | **Tea, coffee, water and soft drinks** |  |
| #23 Beef, veal and dishes | #51 Tea, coffee and water |  |
| #24 Lamb and dishes | #58 Soft drinks, not low calorie |  |
| #25 Pork and dishes | #59 Soft drinks, low calorie |  |
| #26 Coated Chicken |  |  |
| #27 Chicken and turkey dishes |  |  |
| #28 Liver, products and dishes |  |  |
| #29 Burgers and kebabs |  |  |
| #30 Sausages |  |  |
| #31 Meat pies and pastries |  |  |
| #32 Other meat and meat products |  |  |
| #33 White fish coated or fried |  |  |
| #34 Other white fish, shellfish and fish dishes |  |  |
| #35 Oily fish |  |  |
| **All meat, and meat dishes (22-32)** |  |  |
| #22 Bacon and ham |  |  |
| #23 Beef, veal and dishes |  |  |
| #24 Lamb and dishes |  |  |
| #25 Pork and dishes |  |  |
| #26 Coated Chicken |  |  |
| #27 Chicken and turkey dishes |  |  |
| #28 Liver, products and dishes |  |  |
| #29 Burgers and kebabs |  |  |
| #30 Sausages |  |  |
| #31 Meat pies and pastries |  |  |
| 32 Other meat and meat products |  |  |
| **All fish and fish dishes (33-35)** |  |  |
| #33 White fish coated or fried |  |  |
| #34 Other white fish, shellfish and fish dishes |  |  |
| #35 Oily fish |  |  |
| **Fruit and vegetables** |  |  |
| #36 Salad and other raw vegetables |  |  |
| #37 Vegetables (not raw) |  |  |
| #40 Fruit |  |  |
| **Salad and other raw vegetables** |  |  |
| #36 Salad and other raw vegetables |  |  |
| **Vegetables (not raw)** |  |  |
| #37 Vegetables (not raw) |  |  |
| **#All potatoes and potato products (38, 39)** |  |  |
| #38 Chips, fried and roast potatoes and potato products |  |  |
| #39 Other potatoes, potato salads and dishes |  |  |
| **Fruit** |  |  |
| #40 Fruit |  |  |
| **Nuts and seeds** |  |  |
| #56 Nuts and seeds |  |  |

*^*^The number denotes NDNS food or drink group code.*

**Supplementary Table 2. Energy density of excluded minimally processed and ultra-processed food and drink, overall, and within healthy items only.**

| **Energy density of excluded items** | **MPF** | | | **UPF** | | | **P value** |
| --- | --- | --- | --- | --- | --- | --- | --- |
| **kcal per g** | **N** | **Median** | **IQR** | **N** | **Median** | **IQR** |  |
| **All food excluded from analysis** |  |  |  |  |  |  |  |
| All items | 52 | 2.58 | 0.72;3.23 | 767 | 3.40 | 2.31;4.36 | <0.001 |
| Healthy items | 29 | 0.74 | 0.26;2.52 | 258 | 2.30 | 1.46;2.74 | <0.001 |
| **All drink excluded from analysis** |  |  |  |  |  |  |  |
| All items | 0 | NA | NA | 37 | 1.51 | 0.66;2.40 |  |
| Healthy items | 0 | NA | NA | 13 | 0.66 | 0.48;0.84 |  |

*Abbreviations: IQR: interquartile range; MPF: minimally processed food; UPF: ultra-processed food.*

**Supplementary Table 3. Energy density of minimally processed and ultra-processed food by group, overall, and within healthy items only.**

| **By NDNS food group** | **MPF** | | | **UPF** | | |  |
| --- | --- | --- | --- | --- | --- | --- | --- |
| **kcal per 100g** | **N** | **Median** | **IQR** | **N** | **Median** | **IQR** | **P value** |
| **All breakfast cereals** |  |  |  |  |  |  |  |
| All items | 9 | 1.12 | 0.84;1.12 | 86 | 3.70 | 3.56;3.87 | <0.001 |
| Healthy items | 9 | 1.12 | 0.84;1.12 | 59 | 3.70 | 3.48;3.87 | <0.001 |
| High fibre breakfast cereals |  |  |  |  |  |  |  |
| All items | 9 | 1.12 | 0.84;1.12 | 55 | 3.6 | 3.42;3.72 | <0.001 |
| Healthy items | 9 | 1.12 | 0.84;1.12 | 43 | 3.6 | 3.36;3.79 | <0.001 |
| Pasta, rice and other cereals |  |  |  |  |  |  |  |
| All items | 67 | 3.28 | 1.44;3.55 | 83 | 2.02 | 1.36;2.57 | 0.001 |
| Healthy items | 66 | 3.29 | 1.46;3.55 | 65 | 1.74 | 1.23;2.26 | <0.001 |
| **Fruit and vegetables** |  |  |  |  |  |  |  |
| All items | 422 | 0.38 | 0.22;0.87 | 103 | 1.28 | 0.91;2.09 | <0.001 |
| Healthy items | 375 | 0.33 | 0.21;0.73 | 79 | 1.01 | 0.78;1.51 | <0.001 |
| Salad and other raw vegetables |  |  |  |  |  |  |  |
| All items | 71 | 0.23 | 0.17;0.33 | 15 | 0.98 | 0.94;1.27 | <0.001 |
| Healthy items | 70 | 0.24 | 0.18;0.34 | 14 | 0.98 | 0.96;1.28 | <0.001 |
| Vegetables (not raw) |  |  |  |  |  |  |  |
| All items | 216 | 0.62 | 0.20;1.11 | 80 | 1.42 | 0.91;2.18 | <0.001 |
| Healthy items | 198 | 0.50 | 0.19;0.94 | 62 | 1.12 | 0.78;1.76 | <0.001 |
| Fruit |  |  |  |  |  |  |  |
| All items | 135 | 0.41 | 0.30;0.74 | 8 | 1.99 | 0.78;3.13 | <0.001 |
| Healthy items | 107 | 0.36 | 0.26;0.45 | 3 | 0.77 | 0.77;0.78 | 0.007 |
| **Nuts and seeds** |  |  |  |  |  |  |  |
| All items | 29 | 5.81 | 4.81;6.12 | 5 | 4.97 | 3.50;5.23 | 0.041 |
| Healthy items | 2 | 1.45 | 1.33;1.58 | 0 | NA | NA |  |
| **All potato products** |  |  |  |  |  |  |  |
| All items | 29 | 0.97 | 0.70;1.68 | 32 | 2.13 | 1.60;2.78 | <0.001 |
| Healthy items | 27 | 0.91 | 0.70;1.45 | 29 | 1.9 | 1.46;2.77 | <0.001 |
| Chips, fried and roast potatoes and potato products |  |  |  |  |  |  |  |
| All items | 11 | 1.69 | 1.33;1.92 | 22 | 2.14 | 1.90;2.79 | 0.002 |
| Healthy items | 11 | 1.69 | 1.33;1.92 | 21 | 2.14 | 1.90;2.79 | 0.002 |
| Other potatoes, potato salads and dishes |  |  |  |  |  |  |  |
| All items | 18 | 0.74 | 0.70;0.91 | 10 | 1.43 | 1.16;2.17 | 0.013 |
| Healthy items | 16 | 0.72 | 0.69;0.84 | 8 | 1.30 | 1.08;1.45 | 0.015 |
| **Egg and egg dishes** |  |  |  |  |  |  |  |
| All items | 23 | 1.72 | 1.37;2.01 | 11 | 2.59 | 2.28;2.75 | 0.001 |
| Healthy items | 15 | 1.43 | 1.23;1.62 | 5 | 2.10 | 2.09;2.46 | 0.001 |
| **All meat/fish, and meat/fish dishes** |  |  |  |  |  |  |  |
| All items | 257 | 1.47 | 1.09;1.92 | 291 | 2.12 | 1.41;2.63 | <0.001 |
| Healthy items | 217 | 1.33 | 1.04;1.76 | 169 | 1.59 | 1.23;2.28 | <0.001 |
| **All meat, and meat dishes** |  |  |  |  |  |  |  |
| All items | 189 | 1.65 | 1.14;1.92 | 242 | 2.15 | 1.39;2.68 | <0.001 |
| Healthy items | 160 | 1.47 | 1.12;1.81 | 130 | 1.52 | 1.20;2.36 | 0.010 |
| **All fish and fish dishes** |  |  |  |  |  |  |  |
| All items | 68 | 1.06 | 0.76;1.69 | 49 | 2.04 | 1.47;2.25 | <0.001 |
| Healthy items | 57 | 1.03 | 0.76;1.36 | 39 | 2.04 | 1.47;2.24 | <0.001 |
| Beef, veal and dishes |  |  |  |  |  |  |  |
| All items | 59 | 1.71 | 1.14;1.92 | 33 | 1.19 | 0.95;1.30 | 0.006 |
| Healthy items | 55 | 1.65 | 1.11;1.83 | 29 | 1.19 | 0.95;1.26 | 0.003 |
| Lamb and dishes |  |  |  |  |  |  |  |
| All items | 29 | 1.43 | 1.12;2.13 | 8 | 1.53 | 1.07;1.92 | 0.481 |
| Healthy items | 18 | 1.15 | 1.12;1.40 | 6 | 1.28 | 1.03;1.56 | 0.734 |
| Pork and dishes |  |  |  |  |  |  |  |
| All items | 23 | 1.91 | 1.51;2.65 | 9 | 1.63 | 1.17;2.37 | 0.450 |
| Healthy items | 14 | 1.60 | 1.28;1.84 | 5 | 1.17 | 1.16;1.23 | 0.194 |
| Chicken and turkey dishes |  |  |  |  |  |  |  |
| All items | 51 | 1.48 | 1.09;1.75 | 37 | 1.31 | 1.17;1.55 | 0.806 |
| Healthy items | 50 | 1.42 | 1.09;1.71 | 34 | 1.34 | 1.18;1.66 | 0.993 |
| Liver, products and dishes |  |  |  |  |  |  |  |
| All items | 12 | 1.73 | 1.37;1.85 | 5 | 2.85 | 2.26;2.85 | 0.003 |
| Healthy items | 12 | 1.73 | 1.37;1.85 | 0 | NA | NA |  |
| Other meat and meat products |  |  |  |  |  |  |  |
| All items | 15 | 1.95 | 1.20;2.23 | 20 | 2.62 | 2.05;3.00 | 0.012 |
| Healthy items | 11 | 1.37 | 1.10;1.96 | 5 | 1.45 | 1.41;2.50 | 0.212 |
| Other white fish, shellfish and fish dishes |  |  |  |  |  |  |  |
| All items | 47 | 0.91 | 0.75;1.08 | 18 | 1.24 | 1.14;1.97 | <0.001 |
| Healthy items | 44 | 0.91 | 0.75;1.10 | 15 | 1.24 | 1.15;1.96 | <0.001 |
| Oily fish |  |  |  |  |  |  |  |
| All items | 20 | 2.33 | 1.71;2.48 | 9 | 2.04 | 1.70;3.53 | 0.888 |
| Healthy items | 12 | 1.77 | 1.38;2.21 | 4 | 1.62 | 1.39;1.73 | 0.332 |
| **Puddings, Yogurt, fromage frais and other dairy desserts** |  |  |  |  |  |  |  |
| All items | 15 | 0.77 | 0.62;1.14 | 83 | 1.08 | 0.80;2.10 | 0.011 |
| Healthy items | 13 | 0.77 | 0.62;1.14 | 56 | 0.86 | 0.73;1.08 | 0.119 |
| Yogurt, fromage frais and other dairy desserts |  |  |  |  |  |  |  |
| All items | 13 | 0.77 | 0.60;0.77 | 49 | 0.87 | 0.77;1.38 | 0.035 |
| Healthy items | 11 | 0.75 | 0.60;0.77 | 37 | 0.80 | 0.63;0.96 | 0.060 |

*Abbreviations: IQR: interquartile range; MPF: minimally processed food; UPF: ultra-processed food.*
